# Patterns and predictors of antibiotic use among livestock owners in northeast Madagascar

**DOI:** 10.64898/2026.04.09.26350537

**Authors:** Melody Xiao, Quinn Girard, Michelle Pender, Jean Yves Rabezara, Prisca Rahary, Santatra Randrianarisoa, Fidisoa Rasambainarivo, Olivier Rasolofoniaina, Voahangy Soarimalala, Mark M. Janko, Charles L. Nunn

## Abstract

**Purpose:** Antibiotic use (ABU) is a major driver of antimicrobial resistance (AMR), but ABU patterns are poorly understood in low-income countries where the burden of AMR is great and ABU is insufficiently regulated. Here, we report ABU from ten sites ranging from rural villages to small cities in Madagascar, a country with high AMR levels, and present results from modeling to identify factors that may be associated with ABU in this setting.

**Methods:** We conducted surveys of 290 individuals from ten sites in the SAVA Region of northeast Madagascar to gather data on sociodemographic characteristics, agricultural and animal husbandry practices, recent antibiotic use, the antibiotics that participants recalled using in their lifetimes, and the sources of their antibiotics. Using these data, we conducted statistical analyses with a mixed-effects logistic model to determine which characteristics were associated with recent antibiotic use.

**Results:** Nearly all respondents (N=283, 97.6%) reported ABU in their lifetimes, with amoxicillin being the most widely reported antibiotic (N=255, 90.1% of those reporting ABU). All recalled antibiotics were classified as frontline drugs except for ciprofloxacin. Most respondents who reported antibiotic use also reported obtaining antibiotics without prescriptions from local stores (N=273, 96.5%), while only 52.3% (N=148) reported obtaining antibiotics through a prescriptive route, such as from a health clinic or private doctor. Of the 127 individuals (44.9%) who reported recent ABU, men were found to be significantly less likely to have recently taken antibiotics than women.

**Conclusions:** Our findings provide new insights into ABU in agricultural settings in low-income countries, which have historically been understudied in AMR and pharmacoepidemiologic research. Knowledge of ABU patterns supports understanding of AMR dynamics and AMR control efforts in these contexts, such as interventions on inappropriate antibiotic dispensing.

**Key points:**

- Antibiotic use (ABU) in Madagascar is largely unstudied despite its role in antimicrobial resistance (AMR), which Madagascar faces a high burden of.
- ABU was widespread among livestock owners in northeast Madagascar, with the majority of study participants reporting ABU in their lifetimes and most people reporting ABU also having taken antibiotics in the previous three months.
- Most respondents reported obtaining their antibiotics from non-pharmaceutical stores, indicating high levels of unregulated ABU, though more than half also reported sourcing their antibiotics through prescriptive means (like doctors and health clinics).
- Men were less likely than women to have taken antibiotics in the previous three months.
- These findings support the development of interventions to mitigate the burden of AMR in Madagascar and similar contexts while underscoring the need for more comprehensive research on the drivers and patterns of ABU.

**Plain language summary:** In this study, we provide basic information on antibiotic use (ABU) patterns in Madagascar, a country that experiences high levels of resistance but has been particularly understudied in AMR and pharmacological research. We surveyed 290 farmers with livestock from ten sites across northeast Madagascar about their ABU and found that nearly all study participants (N=283, 97.6%) have used antibiotics in their lifetimes, while a little under half of those who reported ABU also reported using antibiotics in the previous three months (N=127, 44.9%). The most used antibiotic was amoxicillin (N=255, 90.1%). Most people obtained their antibiotics from sources that do not require prescriptions, like general stores, indicating that most ABU is unregulated. Through modeling, we also found that men were less likely than women to have taken antibiotics in the previous three months (OR=0.50, CI 0.30-0.82). These findings help us better understand the dynamics of ABU in low-income countries, which have historically been understudied in AMR and pharmacological research. They also support efforts to mitigate the burden of AMR by revealing ABU dynamics that may contribute to the emergence and spread of AMR, as well as identifying targets for intervention to curb inappropriate ABU.

## 1. Introduction

Antimicrobial resistance (AMR) is one of the greatest health challenges in the world, with nearly 4.71 million deaths associated with AMR in 2021.^1^ Resistance is driven predominantly by inappropriate antibiotic use (ABU).^1,2^ Our understanding of ABU patterns derives primarily from high-income countries (HICs). However, morbidity and mortality associated with AMR is higher in low-income countries (LICs) where populations experience high rates of bacterial infections^3,4^ and access to antibiotics is underregulated,^5,6^ creating an urgent but understudied AMR crisis.

Madagascar is an example of this dynamic. In 2021, Madagascar had the 14^th^ highest age-standardized mortality rate associated with AMR among 204 countries.^7^ Despite this, Madagascar has insufficient AMR and ABU surveillance infrastructure.^8,9^ This gap in monitoring may be especially concerning given Madagascar’s heavy dependence on its agricultural sector. For example, over 85% of the country’s population is involved in agriculture – including animal husbandry – as their main livelihood activity.^10,11^ While the impact of agriculture on AMR is well-studied,^12^ the relationship between animal husbandry practices and ABU is poorly characterized. Animal husbandry in an agricultural context may influence ABU via multiple pathways, such as occupational exposures that increase infection risk^13^ or receiving income that increases access to antibiotics.^14^ Understanding these relationships is important for designing contextually appropriate AMR interventions in agricultural LIC settings.

Here, we present an assessment of ABU patterns in northeast Madagascar. We deployed a survey instrument to characterize the extent of antibiotic use, their sources, and the participants’ animal agricultural practices. We conducted statistical modeling to identify relationships between agricultural practices and ABU use, hypothesizing that participation in different forms of animal husbandry and sales is associated with increased likelihood of ABU. Our findings provide insights into the drivers of ABU in an agricultural LIC context, informing both local intervention strategies and efforts to combat AMR in similar settings globally.

## 2. Materials and methods

### 2.1 Ethics statement

All study participants were 18 years or older at the time of the study and provided informed consent. Study protocols were approved by the Duke University Institutional Review Board (2023-0538 and 2025-0491) and the Malagasy Ethics Panel (N°114-MSANP/SG/AGMED/CERBM).

### 2.2 Study setting and design

We investigated ABU in the SAVA Region of northeast Madagascar. This region is especially reliant on agriculture, with over 90% of residents engaging in smallholder subsistence (i.e. rice) and cash crop (i.e. vanilla) farming and high levels of smallholder animal husbandry.^11,15^ This dependence on farming for food security, livelihood, and health makes the SAVA Region a valuable area in which to study ABU patterns and drivers.

This project was carried out in June and July of 2025 across ten villages and towns around Marojejy National Park in the SAVA Region of Madagascar (*Figure 1*). The sites were selected to reflect a gradient of market activity and urbanicity, based on guidance from Malagasy partners and residents. The city of Sambava, the capital of the SAVA Region, represents the most urban setting. In each location, up to thirty farmers who owned animals living in different households were approached in a central location and recruited to participate in a cross-sectional survey (N=290). Consenting participants were compensated with mobile phone credits.

**Figure 1.**
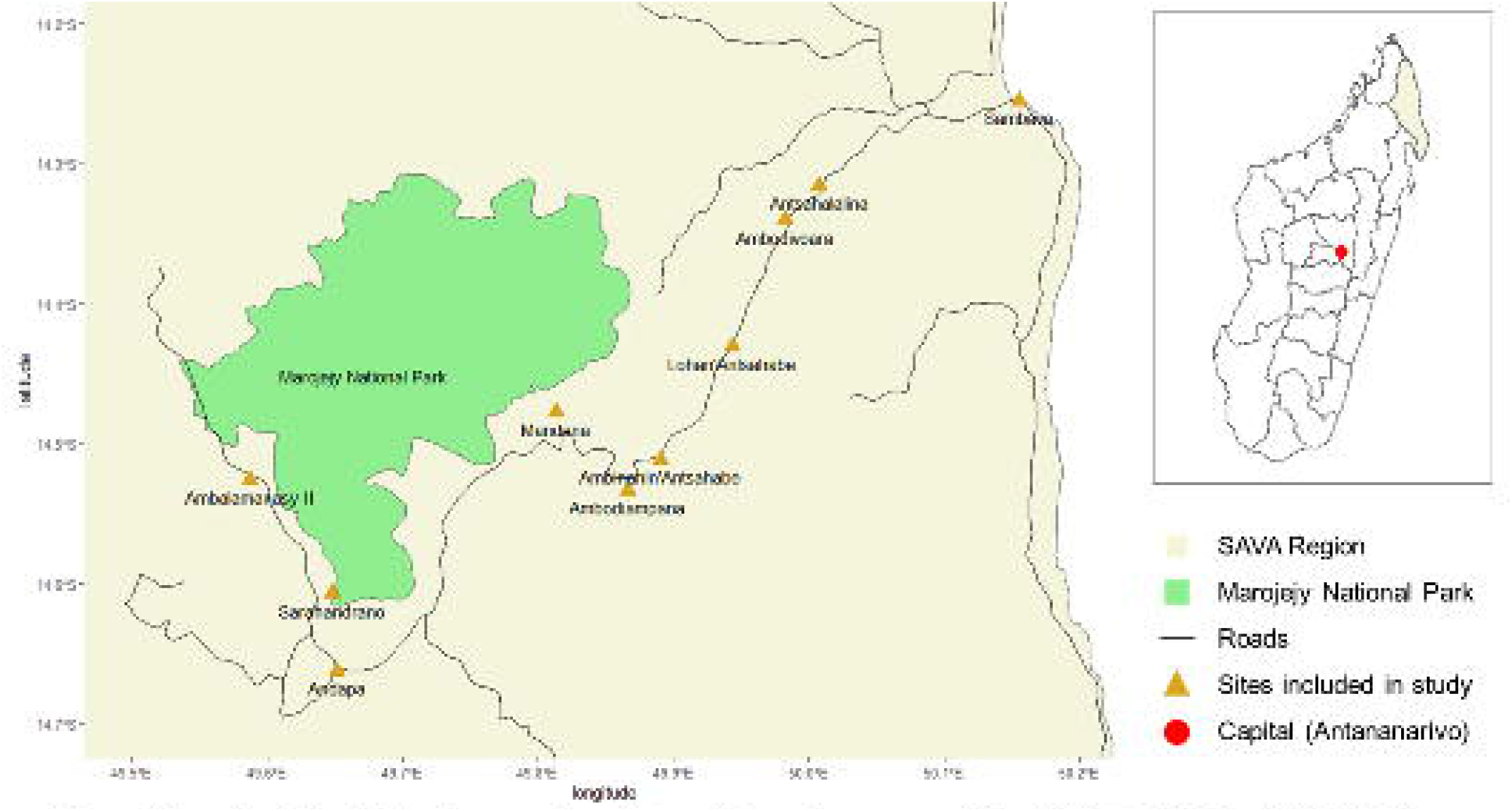
Map of study sites. This study was conducted in ten villages and towns around Marojejy National Park in the SAVA Region of northeast Madagascar. The sites fall along gradients of market participation, level of agriculture, and distance from urban centers.

Survey instruments included demographic characteristics (age and gender), agricultural activities, animal ownership and selling behaviors, and ABU. We focused on rice and vanilla production, as rice is primarily grown for household consumption while vanilla is almost exclusively grown to be sold and indicates participation in the market economy.^11,15^ Participants were also asked how many poultry (i.e., chickens, ducks, and/or geese) and non-poultry livestock (i.e., pigs, goats, and/or zebu/cattle, hereafter *livestock*) they owned, and whether they sold poultry and livestock. The selling of poultry or livestock was also an indicator of market integration.

ABU patterns were determined by (1) asking participants whether they had taken antibiotics in the previous three months and (2) having them identify which antibiotics they remember taking at any point in their lives. To facilitate recall, the survey team showed participants pictures of the pills and packaging of oral antibiotics available at various clinics and stores throughout the SAVA Region. If respondents remembered taking other antibiotics that were not included in the photos, they were asked to name them (*Figure 2*).

**Figure 2.**
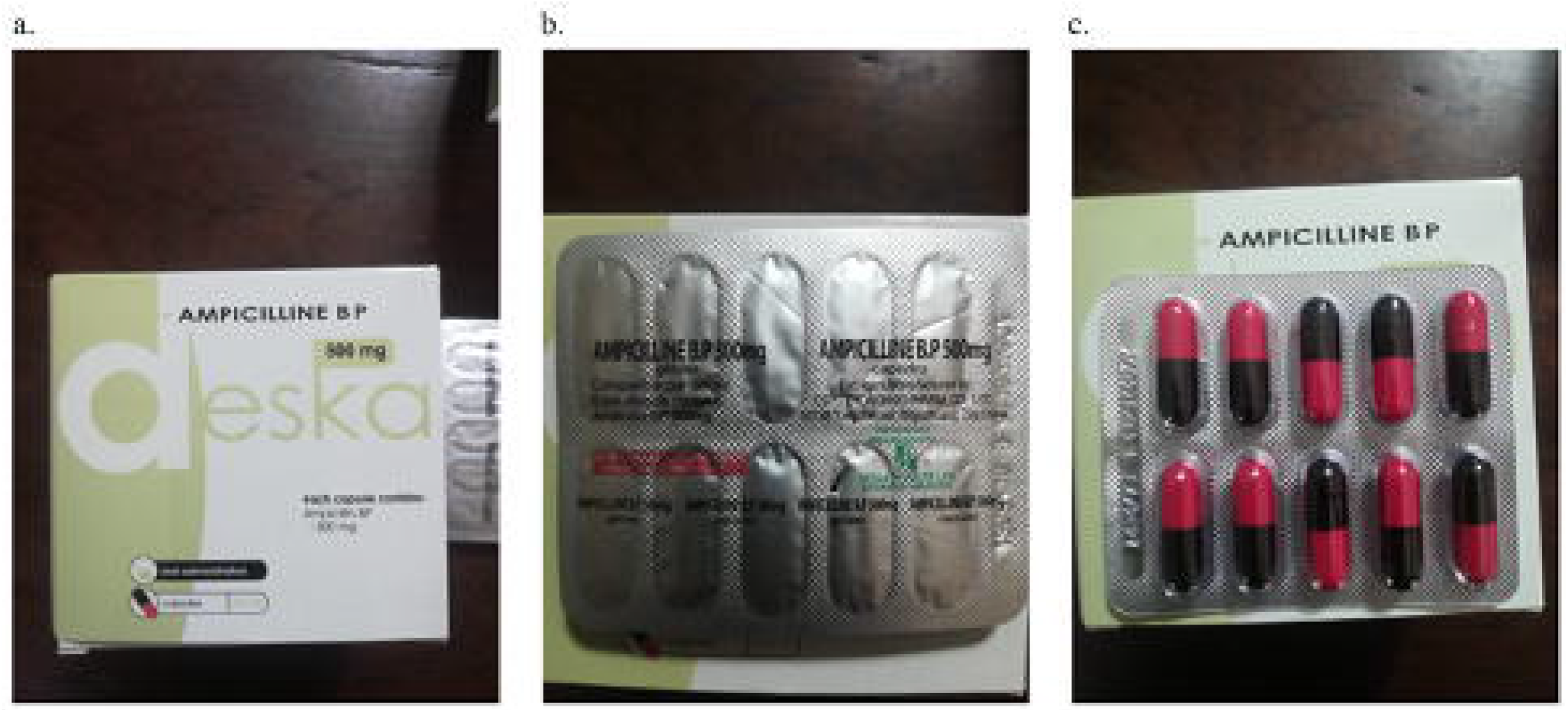
Examples of photos shown to survey participants of antibiotics available at CSBs, pharmacies, and stores in the SAVA Region of Madagascar. Photos courtesy of JYR. (a) Deska brand ampicillin packaging. (b) Front side of ampicillin blister pack. (c) Ampicillin capsules in blister pack.

Participants were also asked to identify the sources of their antibiotics, with options that included private doctors, government-funded primary healthcare clinics (i.e., Centres de Santés de Bases, or CSBs), pharmacies, stores, and friends and family. In this setting, the distinctions between sources are important as they reflect differential access to diagnostics-guided antibiotic prescribing. Obtaining antibiotics from doctors is the most likely method to involve targeted prescriptions based on symptomology consistent with bacterial infections. Meanwhile, while CSBs may be able to diagnose diseases and prescribe antibiotics, they are often understaffed, do not have the capacity to conduct bacteriological testing for more specific antibiotic prescription, and may not have access to more expensive or effective antibiotics.^16^ The use of the term “pharmacy” can refer to one of two sources in Madagascar: a true pharmacy, mostly found in urban areas, that is run by a certified pharmacist and requires prescriptions for drug dispensing; or a “medicament depot,” which sells essential medications in rural or more isolated areas and is approved by the Ministry of Health but often does not require prescriptions to provide antibiotics to patients.^17^ Many small stores and markets also sell antibiotics without prescriptions, allowing for easy access to antibiotics for self-medication without medical guidance.^9^ Lastly, people can obtain antibiotics from friends or family, who may have leftover pills from previously unfinished courses of antibiotics.

### 2.3 Statistical analysis

We computed descriptive statistics to summarize demographics, agricultural practices, animal ownership, animal selling, and ABU in the ten study sites. To assess the impact of demographic and agricultural characteristics on ABU, we fit a mixed-effects logistic regression model with recent antibiotic use as the outcome variable and gender, age, animal ownership, animal selling, and crop growing as covariates. The following levels were set as the reference (baseline) for comparison: women gender, no poultry selling, no livestock selling, no rice growing, and no vanilla growing. Location was included as a random effect to account for the nested study design (study participants in villages/towns).

This model was fitted and analyzed with the *lme4* package^18^ in R version 4.5.2.^19^

## 3. Results

### 3.1 Village and ABU characteristics

We surveyed 290 people across ten sites. Demographic, animal rearing, animal selling, and agricultural characteristics varied across sites. The average age of participants was 40.7 years (ranging from 18 to 88 years), with a roughly even distribution of women and men (51.7 % versus 48.3%). Most participants grew rice (86.9%) and vanilla (70.7%), though the proportion of respondents who participated in growing these crops varied across locations. On average, people tended to own more poultry (14.7 animals) than non-poultry livestock (1.6 animals). A larger proportion of participants also reported selling poultry than livestock (49.0% versus 9.0%) (*Table 1*).

**Table 1.** Table of demographic, animal ownership and selling, and agricultural characteristics by site.

|  | Ambalama-<br>nasy II<br>(N = 33) | Ambinanin'<br>Antsahabe<br>(N = 29) | Ambodia-<br>mpana<br>(N = 31) | Ambodivo-<br>ara<br>(N = 21) | Andapa<br>(N = 30) | Antsahala-<br>lina<br>(N = 30) | Lohan'<br>Antsahabe<br>(N = 30) | Mandena<br>(N = 28) | Sambava<br>(N = 30) | Sarahand-<br>rano<br>(N = 28) | TOTAL<br>(N = 290) |
| --- | --- | --- | --- | --- | --- | --- | --- | --- | --- | --- | --- |
| age in years<br>mean<br>(range) | 48.3<br>(20 – 75) | 44.1<br>(19 – 71) | 36.1<br>(18 – 76) | 40.7<br>(25 – 75) | 42.2<br>(18 – 70) | 39.6<br>(18 – 79) | 37.0<br>(20 – 73) | 41.5<br>(18 – 88) | 41.7<br>(18 – 66) | 34.8<br>(19 – 68) | 40.7<br>(18 – 88) |
| gender<br>% women<br>/ % men | 78.8 / 21.2 | 34.5 / 65.5 | 35.5 / 64.5 | 57.1 / 42.9 | 50.0 / 50.0 | 46.7 / 53.3 | 46.7 / 53.3 | 60.7 / 39.3 | 60.0 / 40.0 | 46.4 / 53.6 | 51.7 / 48.3 |
| grow rice<br>% growers | 87.9 | 86.2 | 93.5 | 85.7 | 76.7 | 90.0 | 100.0 | 100.0 | 56.7 | 92.9 | 86.9 |
| grow vanilla<br>% growers | 63.6 | 86.2 | 77.4 | 76.2 | 30.0 | 66.7 | 93.3 | 89.3 | 56.7 | 71.4 | 70.7 |
| livestock<br>owned<br>mean<br>(range) | 1.8<br>(0 – 20) | 1.2<br>(0 – 6) | 2.2<br>(0 – 10) | 2.1<br>(0 – 7) | 1.7<br>(0 – 6) | 2.3<br>(0 – 15) | 1.9<br>(0 – 7) | 0.6<br>(0 – 5) | 1.8<br>(0 – 17) | 0.8<br>(0 – 3) | 1.6<br>(0 – 20) |
| poultry<br>owned<br>mean<br>(range) | 19.9<br>(0 – 60) | 8.4<br>(0 – 53) | 20.9<br>(0 – 109) | 17.1<br>(0 – 91) | 13.6<br>(0 – 36) | 9.8<br>(0 – 30) | 11.9<br>(0 – 40) | 14.5<br>(3 – 55) | 20.1<br>(0 – 228) | 9.8<br>(0 – 30) | 14.7<br>(0 – 228) |
| sell<br>livestock<br>% sellers | 3.0 | 10.3 | 6.5 | 23.8 | 16.7 | 13.3 | 3.3 | 3.6 | 13.3 | 0.0 | 9.0 |
| sell poultry<br>% sellers | 63.6 | 31.0 | 54.8 | 38.1 | 56.7 | 33.3 | 43.3 | 64.3 | 50.0 | 50.0 | 49.0 |

Nearly every survey participant reported taking an oral antibiotic in their lifetime (N=283, 97.6%) (*Figure 3a*). The seven respondents who reported no antibiotic use in their lifetimes were from three locations (Andapa, Antsahalalina, and Sambava), two of which are among the largest sites included in this study (Andapa and Sambava, both of which are cities). The most commonly reported antibiotic was amoxicillin (N=255, 87.9%), followed by metronidazole (N=131, 45.2%), co-trimoxazole/sulfamethoxazole and trimethoprim (N=120, 41.4%), ampicillin (N=80, 27.6%), ciprofloxacin (N=56, 19.3%), tetracycline (N=33, 11.4%), cloxacillin (N=9, 3.1%), doxycycline (N=5, 1.7%), and Fleming (i.e., amoxicillin and clavulanic acid, N=1, 0.3%). Amoxicillin, ciprofloxacin, co-trimoxazole, metronidazole, and tetracycline use were reported in all villages, with amoxicillin being the most-used antibiotic at every location (*Supplementary Table 1*). Most respondents (N=218, 75.2%) reported using more than one antibiotic in their lifetimes, and 127 respondents (44.9%) reported ABU in the last three months.

**Figure 3.**
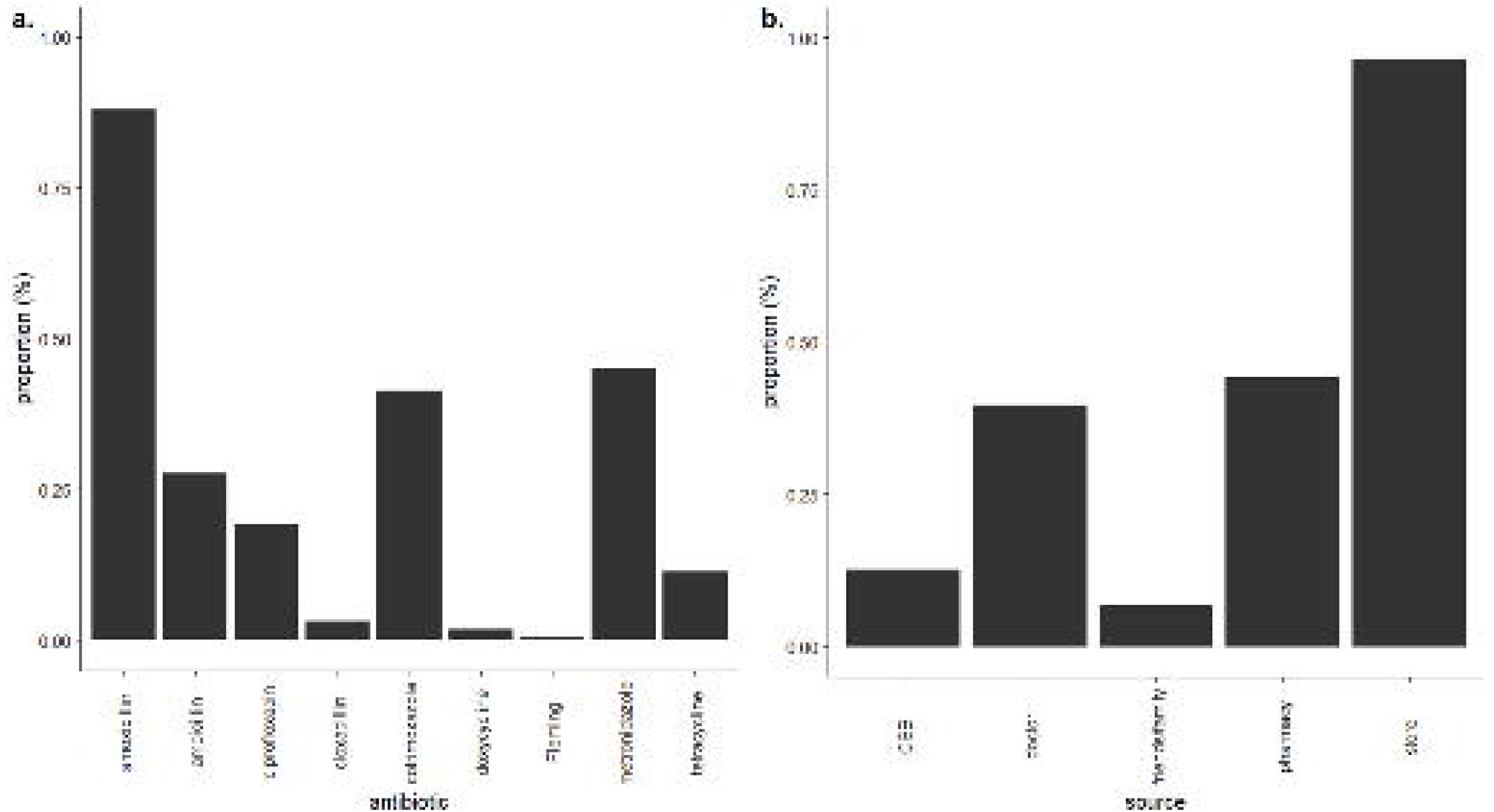
Bar plots of ABU and antibiotic sources. (a) Percent of respondents reporting use of each antibiotic. (b) Percent of respondents reporting antibiotic acquisition from each source.

All survey participants who reported antibiotic use also identified where they acquired antibiotics (*Figure 3b*) with variation across sites (*Supplementary Table 2*). Most participants reported obtaining antibiotics from a store without a prescription (N=273, 96.5% of participants reporting ABU). Nearly half obtained antibiotics from pharmacies (N=125, 44.2%), 39.6% (N=112) obtained antibiotics from a private doctor, 12.7% (N=36) acquired antibiotics from a CSB, and 6.7% (N=19) obtained their antibiotics from friends or family.

### 3.2 Modeling ABU factors

Results from the mixed-effects logistic model show statistical relationships between demographic characteristics and ABU in the previous three months, but no statistically significant relationships between any agricultural or animal ownership characteristics and recent ABU when controlling for variation among sites. Men were less likely than women to have taken antibiotics in the three months prior to the survey (OR = 0.5, 95% CI 0.30-0.82), while age was positively associated with recent ABU (OR = 1.02, 95% CI 1.00-1.03) (*Table 2*).

**Table 2.** Effects of predictors on ABU in the last three months, presented as odds ratios and confidence intervals of logistic mixed-effects model assessing the relationship between recent ABU and fixed effects (variables) while controlling for sites as random effects. * = p < 0.05

| *Variable* | *Coefficient as odds ratio [confidence interval]* |
| --- | --- |
| Men | 0.50 [0.30, 0.82]* |
| Age | 1.02 [1.00, 1.03] |
| Number of poultry owned | 1.01 [0.99, 1.02] |
| Number of livestock owned | 0.99 [0.89, 1.11] |
| Poultry selling | 0.70 [0.42, 1.17] |
| Livestock selling | 0.49 [0.18, 1.30] |
| Rice growing | 0.76 [0.34, 1.71] |
| Vanilla growing | 1.52 [0.83, 2.78] |

## 4. Discussion

As the threat of AMR continues to grow, it becomes increasingly important to characterize patterns of ABU. However, ABU patterns in low-income countries remain poorly described,^5^ including in Madagascar.^9^ This study contributes to understanding ABU in Madagascar with a focus on an understudied rural setting. Among 290 survey participants across ten sites, 283 people reported ABU in their lifetimes, with amoxicillin being the most used antibiotic. Most respondents who took antibiotics reported buying them without prescriptions from local stores. Men were less likely to have taken antibiotics in the previous three months than women, and older respondents were marginally more likely to have used antibiotics. No agricultural, animal ownership, or animal selling characteristics were associated with recent ABU. We review the implications of these findings in the context of other evidence below.

Global ABU surveillance has found that LICs are experiencing dramatic increases in rates of antibiotic consumption and high levels of inappropriate use due to a combination of economic development, irrational prescribing, and unregulated over-the-counter sales.^6,20^ Despite these findings, people living in LICs still face challenges in accessing second-line, broad-spectrum, and last-resort antibiotics, limiting their options for effective treatment when resistance to frontline antibiotics causes them to fail.^4–6,20^ Our findings are consistent with these global patterns; while almost all respondents reported antibiotic use in their lifetimes, all named antibiotics except ciprofloxacin are classified as Access, or frontline antibiotics, in the WHO AWaRe system.^21^ Ciprofloxacin was the only named antibiotic classified as Watch, or broader-spectrum second-line antibiotics, and no Reserve – or last-resort – antibiotics were reported.^21^ This lack of second-line and last-resort antibiotics and near-total reliance on frontline antibiotics, despite the high levels of serious bacterial infections in Madagascar,^22^ indicates barriers to accessing these more costly and specialized drugs.

The finding that most antibiotic users acquired them from stores rather than healthcare facilities is also consistent with global patterns and a cause for concern. Antibiotics in low- and middle-income countries are often acquired without prescriptions due to limited healthcare access and lax drug regulations, leading to high levels of self-medication and inappropriate ABU, including incomplete courses of antibiotics and subtherapeutic dosing.^6,14^ In turn, these apply a selective pressure favoring resistance and lead to higher rates of AMR,^2^ which further increases the burden of infectious disease and limits the options that people have for treatment,^5^ emphasizing the need for interventions focused on antibiotic dispensing. Thus, the finding that people are acquiring antibiotics through non-prescriptive routes in rural Madagascar is an important consideration for infectious disease control in this region.

Our results also contribute to knowledge on drivers of ABU in low-income settings more generally. While our modeling did not find strong evidence of a relationship between ABU and agricultural activity, animal ownership, or animal selling, we found a negative association with being a man and a marginally positive association with age. Global analyses show that women are more likely to receive antibiotic prescriptions in their lifetimes,^23^ possibly due to women seeking out medical care more than men^24,25^ and being more vulnerable to bacterial infections.^26^ However, these analyses were almost entirely based on high-income countries,^23^ which have very different biomedical systems, public health infrastructure, and cultures of health behavior compared to LICs. More work is needed to understand ABU in LIC contexts, especially when the goal is to intervene on inappropriate ABU and mitigate the burden of AMR.

Our study had several limitations worth noting. Firstly, the sample size of this survey (290 people from ten locations) was small and only included livestock owners, and may not be representative of ABU patterns in the whole SAVA Region; however, it should be noted that most households in the SAVA Region do own livestock,^11^ thus animal owners may still account for a wide range of types of antibiotic users. Even so, we tried to account select villages spanning a range of agricultural activity and urbanicity to capture more variation. Secondly, we did not collect ABU timing data (i.e., how long ago ABU occurred and how long the antibiotics were taken) and collapsed ABU into one question. We also did not collect information on participants’ ABU decision-making, such as the conditions treated, the reasons for using specific antibiotics, the dosage, and whether they used them correctly. Thirdly, the variety of drug appearances and packaging may lead to misidentification of antibiotics. Lastly, our survey was conducted cross-sectionally and so modeling of recent ABU only included the three months prior to survey administration, which is during the cool and dry season in Madagascar. ABU patterns may differ depending on the time of year. Additional research on knowledge, attitudes, and practices surrounding ABU would greatly advance research on ABU in LICs.

In conclusion, this study of ABU in the SAVA Region of Madagascar – a tropical agricultural area in an LIC that has generally been understudied in AMR research – provides foundational information on ABU and antibiotic acquisition patterns and identifies individual-level characteristics associated with ABU. These findings support further, more extensive work on ABU in LICs, which will in turn aid AMR control efforts in these contexts.

## Data Availability

Deidentified data may be made available upon request due to privacy and ethical concerns.

## Funding

Funding was provided by the joint NIH-NSF-NIFA Ecology and Evolution of Infectious Disease Program (DEB-2308460) and a pilot grant from Duke/Duke-NUS awarded to MMJ and CLN (Duke/Duke-NUS/RECA(Pilot)/2024/0072). The doctoral work of MX is funded by the Robert Wood Johnson Foundation through the Health Policy Research Scholars Program.

## Declaration of competing interests

The authors declare no conflicts of interest.

## Data sharing and availability

Deidentified data may be made available upon request due to privacy and ethical concerns.

## Acknowledgements

We sincerely thank all survey participants for their invaluable contributions to this study. We also thank everyone who aided in data collection in Madagascar, including the Duke Lemur Center for their logistical support. Lastly, we thank the Malagasy Ethics Panel for their permission to conduct this research.

## Notes

### Competing Interest Statement

The authors have declared no competing interest.

### Author Declarations

The IRB of Duke University gave ethical approval for this work (2023-0538 and 2025-0491). The Malagasy Ethics Panel gave ethical approval for this work (No. 114-MSANP/SG/AGMED/CERBM).

