## Supplemental Table 1 for "Patterns and predictors of antibiotic use among livestock owners in northeast Madagascar"

|  | Ambalama-  nasy II  (N = 33) | Ambinanin’  Antsahabe  (N = 29) | Ambodia-  mpana  (N = 31) | Ambodivo-  ara  (N = 21) | Andapa  (N = 26) | Antsahala-  lina  (N = 28) | Lohan’  Antsahabe  (N = 30) | Mandena  (N = 28) | Sambava  (N = 29) | Sarahand-  rano  (N = 28) | TOTAL  (N = 283) |
| --- | --- | --- | --- | --- | --- | --- | --- | --- | --- | --- | --- |
| amoxicillin | 32 (97.0) | 28 (96.6) | 28 (90.3) | 19 (90.5) | 20 (76.9) | 25 (89.3) | 28 (93.3) | 26 (92.9) | 27 (93.1) | 22 (78.6) | 255 (87.9) |
| ampicillin | 7 (21.2) | 6 (20.7) | 13 (41.9) | 0 (0.0) | 15 (57.7) | 11 (39.3) | 8 (26.7) | 7 (0.25) | 3 (10.3) | 10 (35.7) | 80 (27.6) |
| ciprofloxacin | 13 (39.4) | 5 (17.2) | 3 (9.7) | 4 (19.0) | 3 (11.5) | 4 (14.3) | 5 (16.7) | 6 (21.4) | 8 (27.6) | 5 (17.9) | 56 (19.3) |
| cloxacillin | 4 (12.1) | 0 (0.0) | 0 (0.0) | 1 (4.8) | 2 (7.7) | 0 (0.0) | 0 (0.0) | 0 (0.0) | 2 (6.9) | 0 (0.0) | 9 (3.1) |
| co-trimoxazole | 17 (51.5) | 7 (24.1) | 14 (45.2) | 12 (57.1) | 11 (42.3) | 12 (42.9) | 13 (43.3) | 12 (42.9) | 16 (55.2) | 6 (21.4) | 120 (41.4) |
| doxycycline | 1 (3.0) | 1 (3.4) | 2 (6.5) | 1 (4.8) | 0 (0.0) | 0 (0.0) | 0 (0.0) | 0 (0.0) | 0 (0.0) | 0 (0.0) | 5 (1.7) |
| Fleming | 0 (0.0) | 1 (3.4) | 0 (0.0) | 0 (0.0) | 0 (0.0) | 0 (0.0) | 0 (0.0) | 0 (0.0) | 0 (0.0) | 0 (0.0) | 1 (0.3) |
| metronidazole | 10 (30.3) | 22 (75.9) | 20 (64.5) | 10 (47.6) | 13 (50.0) | 19 (67.9) | 18 (60.0) | 5 (17.9) | 9 (31.0) | 5 (17.9) | 131 (45.2) |
| tetracycline | 4 (12.1) | 6 (20.7) | 1 (3.2) | 3 (14.3) | 6 (23.1) | 4 (14.3) | 4 (13.3) | 2 (7.1) | 2 (6.9) | 1 (3.6) | 33 (11.4) |

*Appendix 1.* Table of antibiotics used by antibiotic and site, among those who reported ABU – number (%)
