## Supplemental Table 2 for "Patterns and predictors of antibiotic use among livestock owners in northeast Madagascar"

|  | Ambalama-  nasy II  (N = 33) | Ambinanin’  Antsahabe  (N = 29) | Ambodia-  mpana  (N = 31) | Ambodivo-  ara  (N = 21) | Andapa  (N = 26) | Antsahala-  lina  (N = 28) | Lohan’  Antsahabe  (N = 30) | Mandena  (N = 28) | Sambava  (N = 29) | Sarahand-  rano  (N = 28) | TOTAL  (N = 283) |
| --- | --- | --- | --- | --- | --- | --- | --- | --- | --- | --- | --- |
| CSB | 1 (3.0) | 0 (0.0) | 5 (16.1) | 1 (4.8) | 7 (26.9) | 1 (3.6) | 9 (30.0) | 0 (0.0) | 6 (20.7) | 6 (21.4) | 36 (12.7) |
| doctor | 29 (87.9) | 0 (0.0) | 1 (3.2) | 0 (0.0) | 14 (53.8) | 3 (10.7) | 3 (10.0) | 27 (96.4) | 12 (41.4) | 23 (82.1) | 112 (39.6) |
| friends or family | 2 (6.1) | 0 (0.0) | 5 (16.1) | 0 (0.0) | 3 (11.5) | 1 (3.6) | 3 (10.0) | 4 (14.3) | 0 (0.0) | 1 (3.6) | 19 (6.7) |
| pharmacy | 29 (87.9) | 0 (0.0) | 7 (22.6) | 0 (0.0) | 26 (100.0) | 7 (25.0) | 4 (13.3) | 16 (57.1) | 13 (44.8) | 23 (82.1) | 125 (44.2) |
| store | 33 (100.0) | 29 (100.0) | 30 (96.8) | 20 (95.2) | 19 (73.1) | 27 (96.4) | 30 (100.0) | 28 (100.0) | 29 (100.0) | 28 (100.0) | 273 (96.5) |

*Appendix 2.* Table of antibiotics used by antibiotic source, among those who reported ABU – number (%)
